# Diagnostic accuracy of deep learning for microsatellite instability prediction in gastric cancer: A systematic review and meta-analysis

**DOI:** 10.64898/2026.10.12.26365351

**Authors:** Yu Du, Hongxia Liao, Jingdong Li, Yinping He, Ji Di

**Author notes:** Corresponding author, (JD).

## Abstract

Deep learning (DL) applied to haematoxylin and eosin (H&E)-stained pathology images can predict microsatellite instability (MSI) in gastric cancer (GC). We evaluated its diagnostic accuracy and discriminative performance. PubMed, Embase, Cochrane Library and Web of Science were searched for studies applying DL to H&E images for MSI prediction in GC. The protocol was prospectively registered in PROSPERO (CRD420261503140). Risk of bias was assessed with QUADAS-2. Pooled sensitivity and specificity were estimated with a bivariate random-effects model, with subgroup analyses by validation strategy and data source; slide-level areas under the curve (AUCs) were pooled after logit transformation. Prespecified scenario analyses assessed robustness. Ten studies were included; seven independent datasets from six studies entered the primary synthesis. Pooled sensitivity, specificity and summary receiver operating characteristic AUC were 0.78 (95% CI 0.63–0.88), 0.85 (95% CI 0.77–0.90) and 0.89 (95% CI 0.86–0.91); nine studies contributed to the secondary AUC synthesis (pooled AUC 0.85, 95% CI 0.80–0.89). Hospital-derived cohorts showed numerically higher sensitivity (0.86) and diagnostic odds ratio (39.8) than TCGA-derived cohorts (0.71 and 11.5), but subgroup differences were limited by the small number of studies and substantial heterogeneity. DL models show good diagnostic accuracy for MSI prediction in GC, but clinical utility still requires confirmation in standardised multicentre, independent external and prospective studies.

## Introduction

Gastric cancer (GC) ranks among the leading causes of cancer incidence and mortality worldwide, with about 968,000 new cases and 660,000 deaths in 2022[1]. Despite advances in precision medicine and systemic treatment, the biological heterogeneity of GC continues to complicate patient stratification and individualised therapy. Molecular classifications based on genomic, epigenetic, and tumour immune microenvironment features indicate that GC is not a single disease entity but comprises several molecular subtypes with distinct biological behaviours and treatment responses [2–3]. Among these, microsatellite instability (MSI), particularly MSI-high (MSI-H) and deficient mismatch repair (dMMR), is a key molecular feature associated with high tumour mutational burden, immune infiltration, and benefit from immunotherapy [4–9]. Accurate identification of MSI status is therefore clinically important for molecular stratification and treatment decisions in GC.

The biological features of MSI-H/dMMR gastric cancer are not confined to the genome. High mutational burden, altered immune microenvironment, and specific tumour histology can leave recognisable morphological signals on routine haematoxylin and eosin (H&E)-stained sections [10–11]. Some MSI-H gastric cancers show richer tumour-infiltrating lymphocytes, mucinous or poorly differentiated morphology, and characteristic tumour site and histological features [12–13]. These observations suggest that routine pathology images may indirectly carry phenotypic information related to genomic instability and immune status. At present, MSI/MMR status in GC is confirmed mainly by polymerase chain reaction (PCR), immunohistochemistry (IHC), and next-generation sequencing (NGS) [14]. These methods form the current clinical basis for MSI assessment but require additional molecular or immunohistochemical testing, which adds burden in tissue specimens, turnaround time, and resource allocation [15–17]. Deriving MSI risk cues from routine H&E sections, before confirmatory molecular testing, is therefore a clinically relevant question.

Deep learning (DL) offers a new technical route to this problem. Unlike traditional machine learning methods that rely on manually defined features, DL models learn multi-level visual representations of tissue architecture, cell morphology, and spatial relationships directly from digital pathology images, and thereby establish statistical associations between image features and MSI status [18]. Kather et al. first showed that a DL model can predict MSI status directly from H&E images of gastrointestinal tumours, suggesting that MSI-related information not reliably identified by the human eye may exist in tissue morphology [19]. Recent multicentre pathology AI studies further indicate that differences in tissue preparation, staining, and digitisation affect model generalisability [20]. Attention-based multiple-instance learning and self-supervised learning have more recently enabled slide-level prediction without fine-grained region-level annotation [21].

Despite this progress, objective evidence on the accuracy of H&E-based deep learning models for MSI prediction in GC remains limited. Existing studies lack a unified evidence-based comparison of model performance across validation strategies, and the heterogeneity arising from different data sources and validation settings is not fully explained.

This study aimed to systematically evaluate the diagnostic accuracy of deep learning models applied to H&E digital pathology images for MSI prediction in GC, to explore sources of heterogeneity through subgroup analyses by data source and validation strategy, and to assess the degree of external validation and features relevant to clinical translation of the existing evidence.

## Materials and methods

### 2.1. Protocol registration and study design

This systematic review and meta-analysis was designed and reported in accordance with the Preferred Reporting Items for Systematic Reviews and Meta-Analyses (PRISMA) guidelines, and the completed checklist is provided as Supporting information. The review protocol was prospectively registered in the International Prospective Register of Systematic Reviews (PROSPERO) under the ID: CRD420261503140.

### 2.2. Search strategy

PubMed, Embase, Cochrane Library, and Web of Science were searched for studies on DL models for diagnosing MSI in gastric cancer, from database inception to 30 June 2026. No restriction was placed on region or publication year. The detailed search procedures and strategies are provided in the Supporting information (S1 Table).

### 2.3. Eligibility criteria

#### 2.3.1. Inclusion criteria

(1) studies of patients with a pathological diagnosis of gastric cancer; (2) studies that fully applied a deep learning model to identify MSI in gastric cancer; (3) original research articles with a prospective or retrospective design, published in English.

#### 2.3.2. Exclusion criteria

(1) other study types, including expert commentary, animal experiments, case reports, meta-analyses, conference abstracts, and reviews; (2) studies lacking a clear reference standard; (3) duplicate publications. This review applied two levels of eligibility. At the study level, studies were included if they concerned a gastric cancer population, used deep learning on H&E pathology images, employed an appropriate reference standard, and reported original research. At the synthesis level, studies contributing to the diagnostic accuracy synthesis had to provide or allow reconstruction of 2×2 contingency tables, those contributing to the AUC synthesis had to report slide-level AUC with 95% CI or standard error, and the remaining included studies were synthesised descriptively.

### 2.4. Data extraction

Two investigators (Du, Liao) independently screened titles and abstracts against the inclusion criteria using EndNote X9 to identify studies for full-text assessment. Data were then extracted independently by the two investigators, with disagreements resolved by discussion. Extracted items included authors, publication year, country/region, study type, total number of cases, data source, model architecture, validation strategy, and 2×2 outcomes, together with AI-specific information on the unit of analysis (patient/slide/tile), whether training and test sets were split at the patient level, data leakage control, model selection procedures, and independence of external validation. Extraction was performed independently and cross-checked, with disagreements resolved by discussion or by a third investigator (Di).

### 2.5. Quality assessment

Quality and risk of bias of the included studies were assessed with QUADAS-2, covering four domains: patient selection, index test, reference standard, and flow and timing. Each domain contains specific signalling questions, judged as “yes”, “no”, or “unclear”, corresponding to low, high, or unclear risk. A domain was rated low risk of bias when all its signalling questions were judged “yes”; a “no” indicated potential bias, and insufficient information was marked “unclear”. Two investigators performed the assessment independently in Review Manager 5.3 and cross-verified the results.

### 2.6. Statistical analysis

A bivariate generalised linear mixed-effects model was used for the meta-analysis, with pooled sensitivity, specificity, positive likelihood ratio, negative likelihood ratio, and diagnostic odds ratio, each with 95% confidence intervals. A summary receiver operating characteristic (SROC) curve was plotted and its area under the curve (AUC) calculated. A secondary synthesis of slide-level AUC was performed for studies reporting this measure, to provide complementary evidence on overall discrimination; individual AUCs were logit-transformed, standard errors were derived from the reported 95% CIs, and random-effects pooling was performed on the logit scale before back-transformation. When a study reported performance for multiple models or test sets, the authors’ prespecified primary model and primary validation cohort were selected; independent validation cohorts from distinct patients were treated as separate datasets, whereas performance estimates from multiple models of the same patient cohort were not treated as independent effect sizes. Given the small number of studies, no formal statistical test for publication bias was performed. Heterogeneity was described with the Cochran Q test and I² statistic, and subgroup analyses by data source and validation strategy were specified to explore potential sources of heterogeneity. All analyses were performed in Stata 17, with p < 0.05 considered statistically significant.

### 2.7. Ethics statement

This study used only published aggregate data and did not involve direct recruitment of human participants. Ethical approval and informed consent were therefore not required.

## Results

### 3.1. Study selection

The searches identified 251 records; after removing 55 duplicates, 14 ineligible records, and 5 records excluded for other reasons, 177 proceeded to title and abstract screening. Twenty-one reports underwent full-text assessment against the prespecified PISO criteria (S2 Table); 11 were excluded (4 non-deep-learning models, 4 with no extractable diagnostic outcome, 2 not using histopathology slides, and 1 not reporting MSI as a separate subgroup). Ten studies were ultimately included in the systematic review (Fig 1).

**Fig 1.**
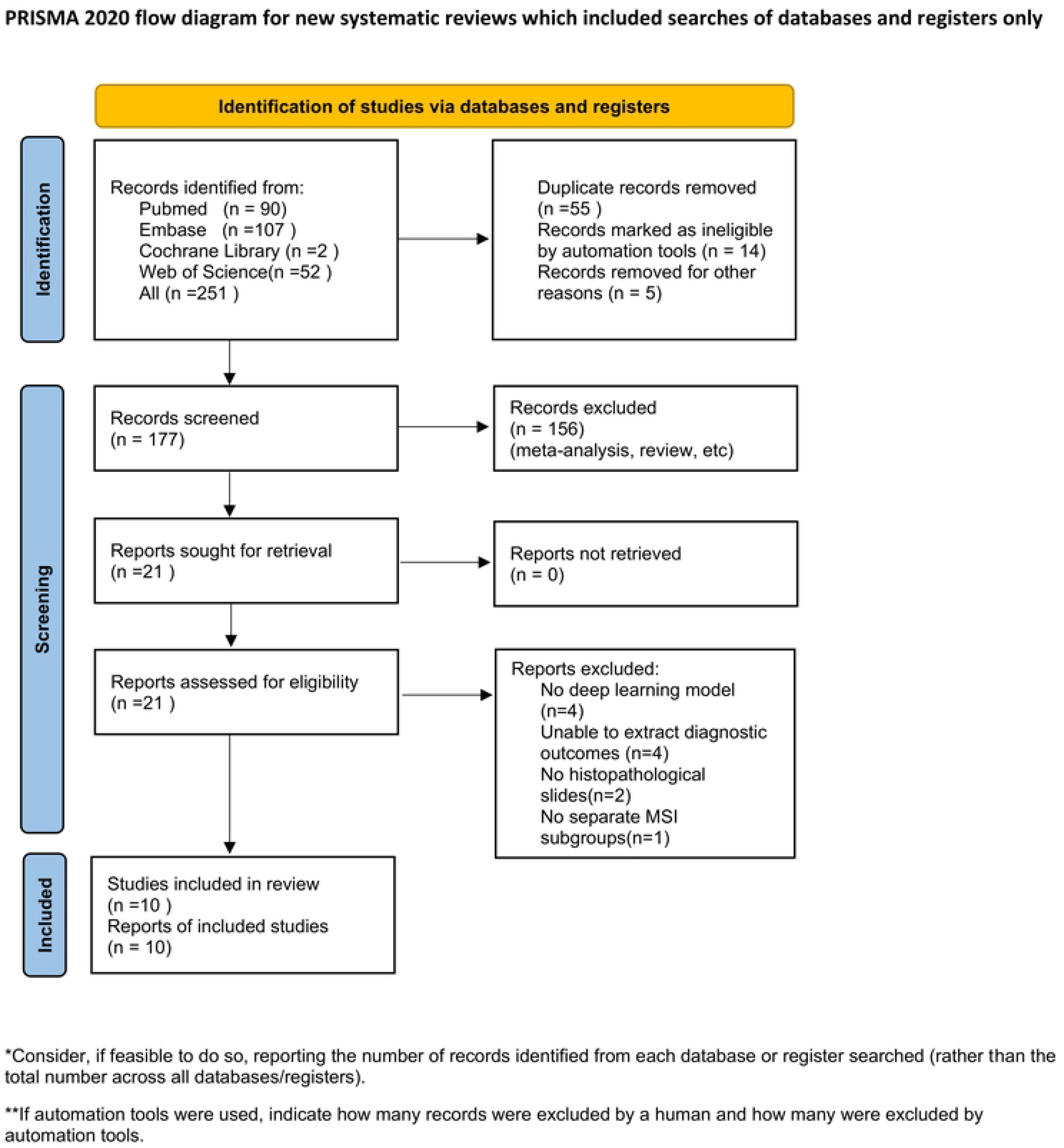
Flow diagram of study selection.

### 3.2. Characteristics of included studies and quality assessment

Ten studies were included [22–31]. All used a deep learning model as the main analytical method and H&E-stained images as the sole model input. The quantitative diagnostic accuracy analysis included seven independent datasets from six studies [22–24,28–30]; Zheng et al.[30] reported two fully independent validation cohorts, a Chinese multicentre hospital cohort and TCGA-STAD, with no overlap in patient source, institution, or country, and these were treated as patient-level independent datasets rather than duplicate counts of one cohort. The secondary AUC analysis included nine studies [22–23,25–31]; Su et al.[24] reported only tile-level AUC, which is not comparable with slide-level AUC and was therefore excluded from pooling. Four studies originated from China [22,24,27,30], two from Korea [23,29], three were international multicentre studies [25,26,31], and one from Australia [28]. Most models used convolutional neural networks [22–25,28,29]; more recent studies introduced attention-based multiple-instance learning [27], self-supervised contrastive learning [26], a StyleGAN-based interpretable framework [30], and Bayesian deep Gaussian processes [31]. Three studies used TCGA-STAD alone as the gastric cancer cohort [27–29], three combined TCGA with hospital or multicentre cohorts [22,23,30], three used international multicentre cohorts [25,26,31], and one used a single-centre hospital cohort [24]. In subgroup analyses, each study was assigned to the hospital or TCGA stratum according to the actual source of its retained AUC data point. Six studies included external or independent validation cohorts [22,23,25,26,30,31], and four used internal validation only [24,27–29]. Details are provided in S3 Table. The certainty of evidence for the main diagnostic outcomes was assessed with the GRADE approach for diagnostic test accuracy (S4 Table), rated as high, moderate, low, or very low across five domains: risk of bias, indirectness, inconsistency, imprecision, and publication bias. All included studies were retrospective. The risk-of-bias and applicability assessment is shown in Fig 2.

**Fig 2.**
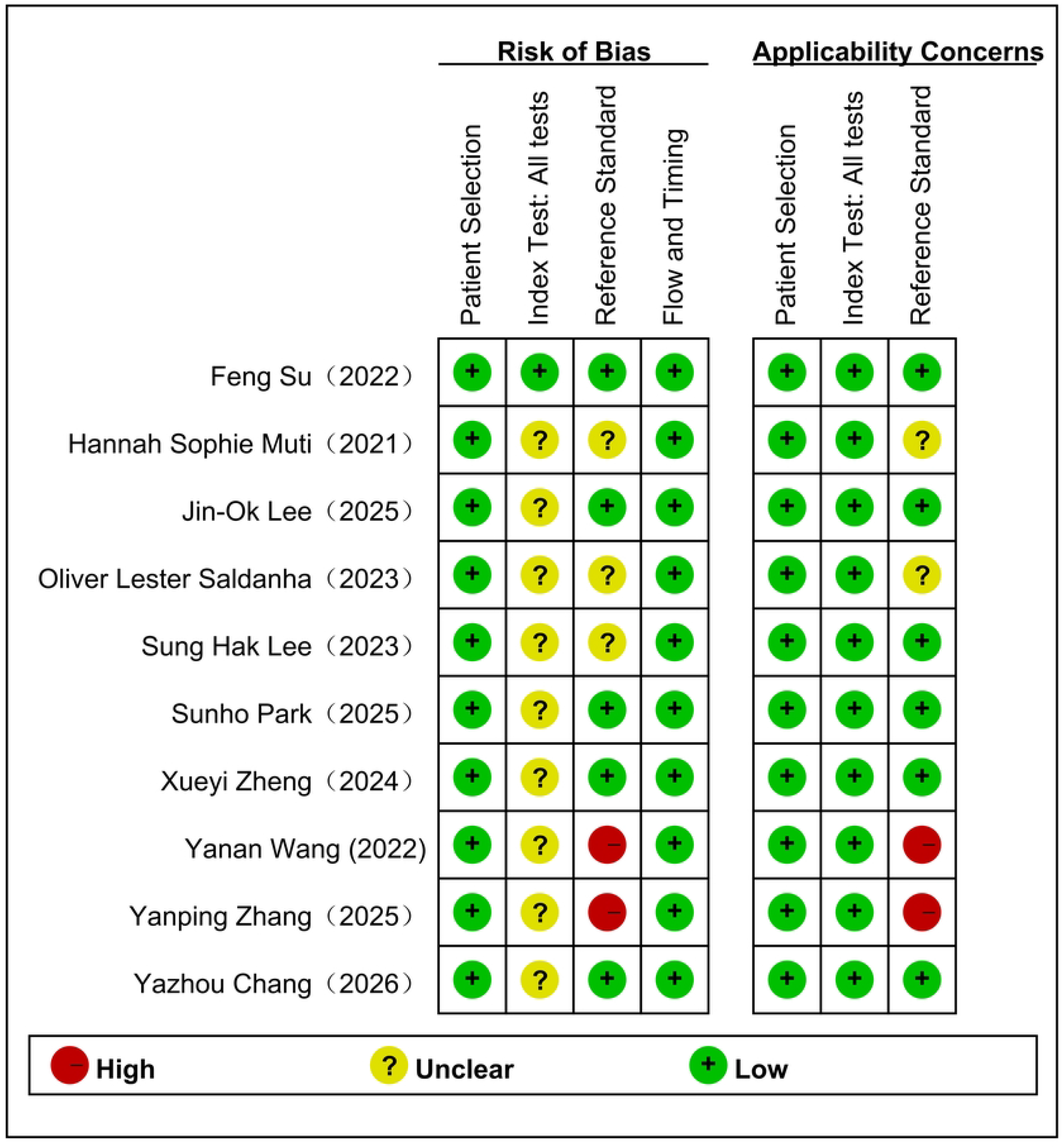
Quality assessment of the included studies.

### 3.3. Meta-analysis of diagnostic accuracy

Seven independent validation datasets from six studies entered the primary diagnostic accuracy meta-analysis, and nine studies reporting comparable slide-level AUC entered the secondary AUC synthesis. The pooled sensitivity was 0.78 (95% CI 0.63–0.88) and the pooled specificity 0.85 (95% CI 0.77–0.90); the positive likelihood ratio (PLR) was 5.18 (95% CI 3.57–7.50), the negative likelihood ratio (NLR) 0.26 (95% CI 0.15–0.44), and the area under the SROC curve 0.89 (95% CI 0.86–0.91) (Fig 3). The secondary AUC meta-analysis yielded a pooled AUC of 0.85 (95% CI 0.80–0.89) (Fig 4), indicating good overall discrimination of MSI status in gastric cancer (details in S1 Fig).

**Fig 3.**
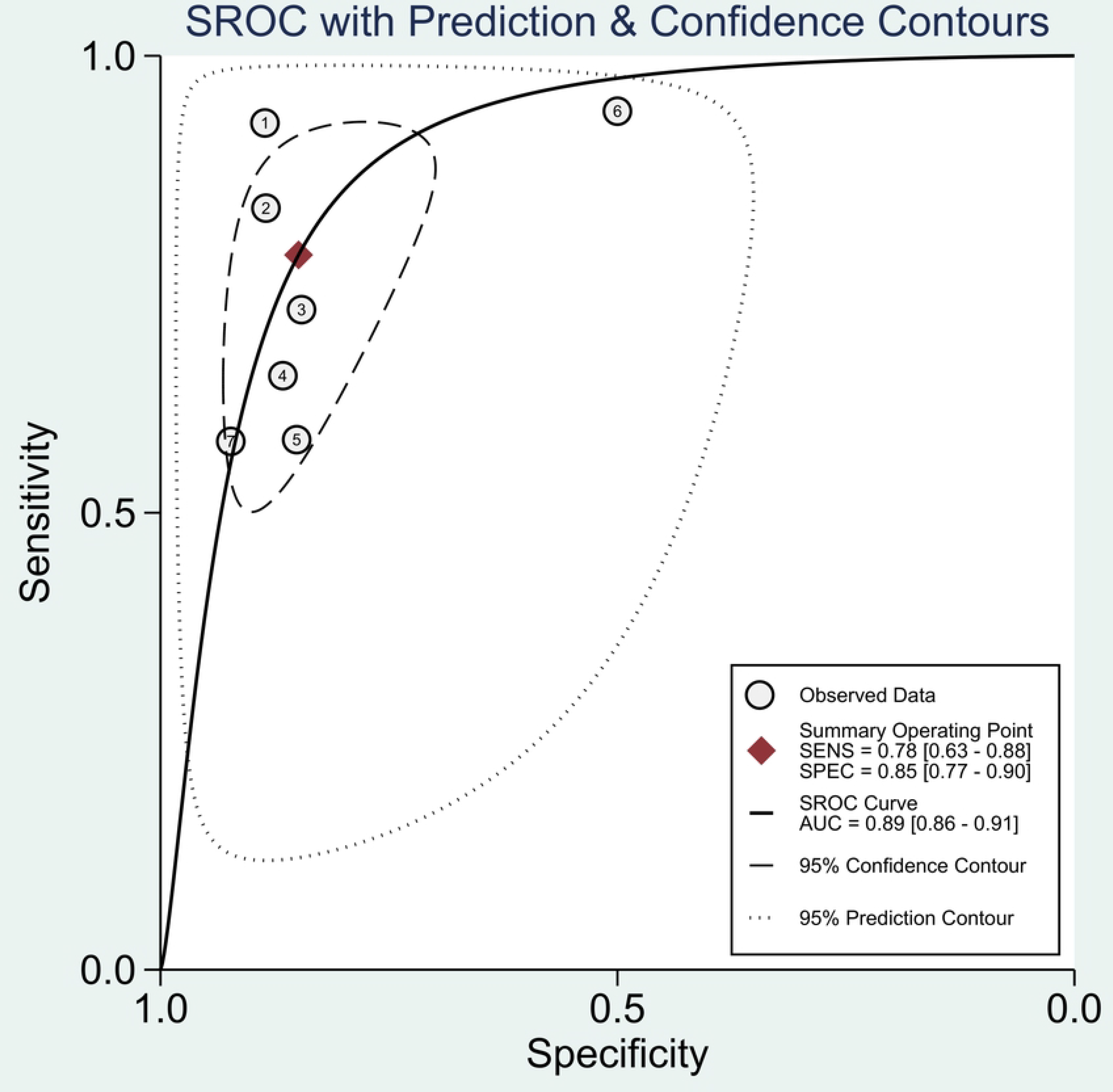
SROC curve for MSI prediction in gastric cancer.

**Fig 4.**
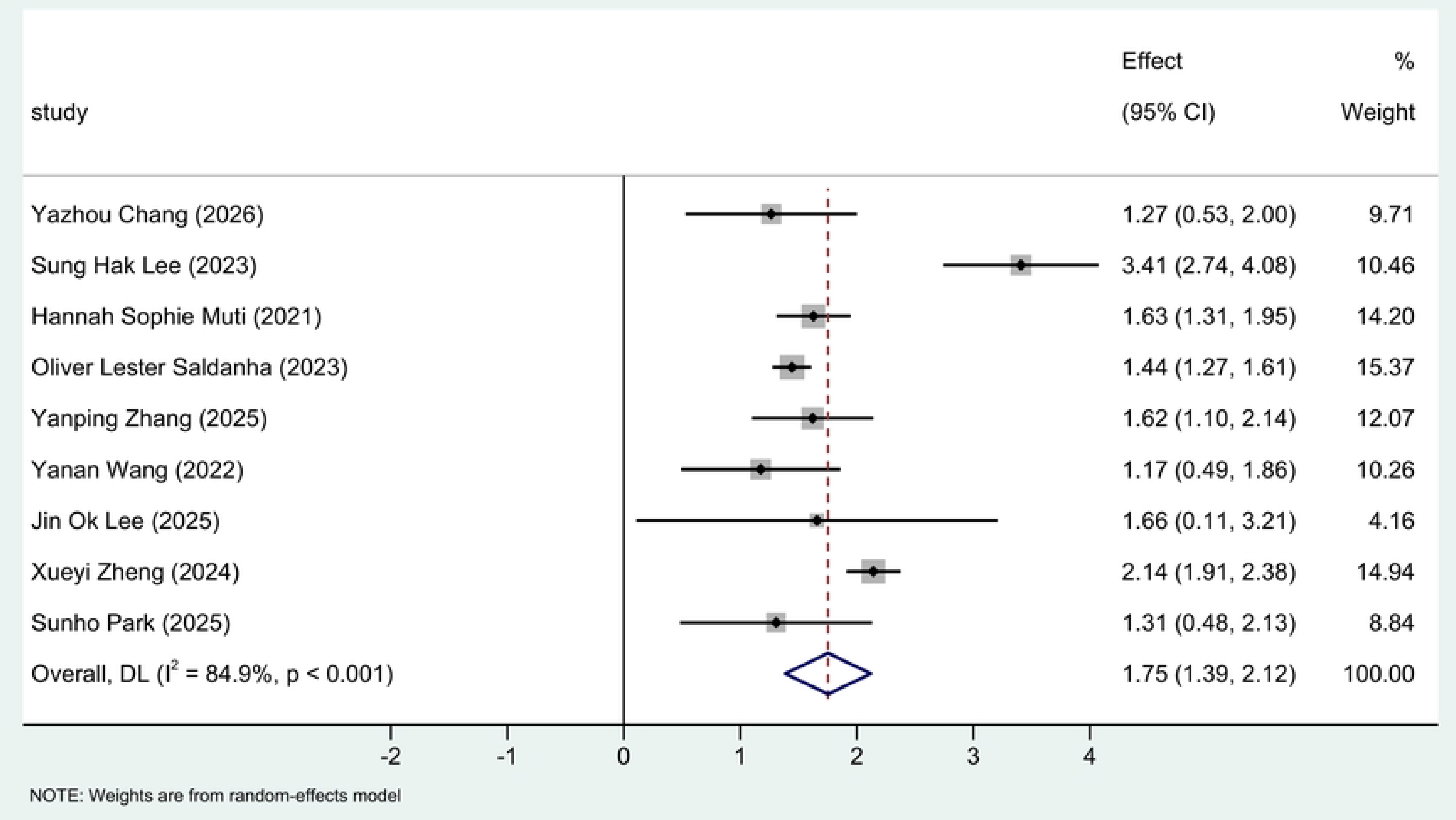
Forest plot of pooled logit-AUC.

### 3.4. Subgroup analyses

Given the substantial heterogeneity in the pooled results, exploratory subgroup analyses were used to explore its sources (Tables 1 and 2). Validation strategy and dataset source were used as stratification variables, and between-group differences in the AUC subgroup were assessed with the Q test.

**Table 1.** Subgroup analyses of pooled diagnostic accuracy of deep learning models for MSI prediction in gastric cancer.

| Subgroup | No. of datasets | Pooled sensitivity (95% CI) | Pooled specificity (95% CI) | DOR (95% CI) | I<sup>2</sup> for sensitivity (%) | I<sup>2</sup> for specificity (%) |
| --- | --- | --- | --- | --- | --- | --- |
| Overall | 7 | 0.78 (0.63–0.88) | 0.85 (0.77–0.90) | 19.9 (8.1–50.0) | 79.8 | 81.6 |
| **Validation strategy** |  |  |  |  |  |  |
| External | 3 | 0.80 (0.59–0.91) | 0.87 (0.84–0.89) | 25.7 | 73.1 | 18.9 |
| Internal | 4 | 0.77 (0.54–0.90) | 0.82 (0.65–0.92) | 14.9 | 83.3 | 85.5 |
| **Dataset source** |  |  |  |  |  |  |
| Hospital | 3 | 0.86 (0.73–0.93) | 0.87 (0.83–0.90) | 39.8 | 44.2 | 13.4 |
| TCGA | 4 | 0.71 (0.51–0.86) | 0.82 (0.65–0.92) | 11.5 | 83.8 | 89.1 |

Corresponding subgroup forest plots are provided in the Supporting information (S2 and S3 Figs).

**Table 2.** Subgroup analyses of pooled discriminative ability (AUC) of deep learning models for MSI prediction in gastric cancer.

| Subgroup | No. of studies | Pooled AUC (95% CI) | I<sup>2</sup> (%) |
| --- | --- | --- | --- |
| Overall | 9 | 0.85 (0.80–0.89) | 84.9 |
| **Validation strategy** |  |  |  |
| External | 5 | 0.87 (0.79–0.93) | 92.0 |
| Internal | 4 | 0.83 (0.79–0.86) | 0.0 |
| **Dataset source** |  |  |  |
| Hospital | 5 | 0.88 (0.79–0.93) | 85.0 |
| TCGA | 4 | 0.81 (0.79–0.84) | 0.0 |

Pooled AUC was back-transformed (invlogit) from logit-transformed random-effects meta-analysis of nine studies reporting slide-level AUC. Corresponding subgroup forest plots are provided in the Supporting information (S4 and S5 Figs).

### 3.5. Robustness analyses

Two prespecified scenario analyses assessed the robustness of the main results to data independence and validation strategy. First, to examine the effect of potential patient overlap in TCGA-STAD and its derivative datasets, studies using TCGA as the data source were excluded; the remaining hospital cohorts showed pooled sensitivity of 0.86 (95% CI 0.73–0.93) and specificity of 0.87 (95% CI 0.83–0.90), consistent in direction with the full analysis (0.78/0.85), and the secondary AUC synthesis excluding TCGA-derived studies yielded a pooled AUC of 0.88 (95% CI 0.79–0.93), consistent with 0.85. Second, when only independent external validation datasets were retained, pooled sensitivity was 0.80 (95% CI 0.59–0.91), specificity 0.87 (95% CI 0.84–0.89), and the pooled AUC of external validation cohorts 0.87 (95% CI 0.79–0.93); none of these changed the direction or statistical significance of the pooled estimates (Tables 1 and 2). These analyses indicate that, although few studies remained after removing TCGA-overlapping or non-external-validation data, the results were directionally stable, supporting data source and validation strategy as key contextual factors in interpreting the findings.

## Discussion

### 4.1. Principal findings

This review systematically evaluated the diagnostic performance of deep learning models applied to H&E-stained images for MSI prediction in gastric cancer. The overall results show that deep learning models can extract discriminative information related to MSI status from routine histology, with good overall diagnostic ability, and the AUC analysis provides complementary evidence on overall discrimination. Beyond reporting performance metrics, we examined how performance varies with data source and validation strategy, asking whether models remain stable across different data environments. This matters for clinical translation, because the clinic presents not a single standardised dataset but real-world pathology material that varies in source, case mix, and digitisation workflow.

### 4.2. Interpretation of subgroup findings

Subgroup analyses showed that hospital-derived cohorts performed better than TCGA-derived cohorts on some measures, whereas externally validated cohorts did not clearly underperform internally validated ones. These differences suggest that the generalisability of a deep learning pathology model should be judged against the specific data generation and validation context, rather than the performance of a single model on a single test set. Although TCGA is a shared resource with high accessibility and relatively standardised structure, studies still differ in case selection, WSI quality control, image preprocessing, label definition, and train/test split; sharing a database therefore does not imply a common data generation process, nor consistent model performance. External validation is not by itself sufficient evidence of generalisability; its value depends on how far the validation cohort differs from the development data in patient source, geography, scanner, staining, and case mix [32–33]. We regard the diagnostic performance of pathology models as strongly context-dependent, related not only to the model but also to case selection, data composition, and the specific validation setting. Generalisability therefore requires independent cohorts that are multi-source, cross-institutional, and representative; a model may show different performance structures in different data environments, and hospital real-world cohorts together with independent external data provide complementary evidence on clinical usability.

### 4.3. Comparison with previous reviews

Previous related work pooled pathology images, radiomics, and clinical variables, providing evidence for the overall feasibility of machine-learning-assisted MSI identification [34]. This review narrows the scope to deep learning models on H&E pathology images and applies a diagnostic test accuracy meta-analysis framework to quantify clinically relevant performance, thereby positioning these models for clinical use in gastric cancer MSI identification.

### 4.4. Evidence maturity and clinical translation

Computational pathology in gastric cancer is currently transitioning from model feasibility toward clinical validity; limited data scale, insufficient data diversity, and scarce external validation remain common [35–36]. Larger multicentre studies are beginning to adopt prospective designs and evaluations of model-assisted diagnosis, and some included studies explored interpretability with Grad-CAM or attention visualisation, but reporting of data source, patient splitting, and model development workflow remains insufficiently transparent. The field is moving from algorithmic performance evaluation toward standardised, reproducible, and clinically oriented evidence [37–38]. H&E-based MSI prediction now has the technical basis to enter multicentre, prospective, real-world clinical evaluation, and may become an adjunct linking routine morphological assessment with molecular classification of gastric cancer.

### 4.5. Strengths and limitations

Several limitations should be noted. First, only seven independent cohorts were included, limiting precise assessment of heterogeneity sources and subgroup differences. Some cohorts derived from TCGA or its derivatives; although study-level independence was assessed and scenario robustness analyses performed, overlap at the patient, slide, or feature level cannot be excluded, and this is an important methodological concern for evidence independence (S5 Table). Second, all included studies were retrospective and lacked individual patient-level covariates, limiting analysis of age, tumour site, stage, histological type, and MSI-H prevalence. Finally, we excluded non-English publications, so language bias may be present.

### 4.6. Future directions

Future research should move from retrospective performance demonstration toward prospective clinical validation. Study designs should prespecify patient inclusion criteria, reference standard, model threshold, handling of missing images, and primary clinical endpoints, and embed the model in routine pathology and molecular diagnostic workflows to evaluate its actual effect on testing efficiency, diagnostic consistency, molecular testing resource allocation, and patient management. Improving generalisability through multicentre training and cross-site validation should take priority over incremental gains in internal validation performance.

## Conclusions

Current evidence indicates that deep learning models applied to H&E pathology images can identify morphological information related to MSI status in gastric cancer, with good overall discriminative performance. However, the evidence derives mainly from retrospective studies with marked heterogeneity in data source and validation strategy; clinical utility therefore still requires confirmation in standardised multicentre, independent external, and prospective studies.

## Acknowledgments

The authors thank the developers of the publicly available datasets and code used in the included primary studies, whose work made this secondary analysis possible.

## Supporting information

**S1 Checklist. PRISMA 2020 checklist for this systematic review.**

**S1 Text. Review protocol registered in PROSPERO (CRD420261503140).**

**S1 Table. Full electronic search strategies for PubMed, Embase, Cochrane Library and Web of Science, with the number of records retrieved per database.**

**S2 Table. List of all records screened at full-text assessment with reasons for exclusion (PISO criteria).**

**S3 Table. Characteristics of the included studies, including model architecture, data source, validation strategy, unit of analysis and reference standard.**

**S4 Table. GRADE assessment of certainty of evidence for the main diagnostic outcomes across five domains.**

**S5 Table. Assessment of potential cohort overlap between the included studies.**

**S1 Fig. Diagnostic accuracy estimates of the individual included datasets.**

**S2 Fig. Subgroup forest plot of pooled sensitivity and specificity by validation strategy.**

**S3 Fig. Subgroup forest plot of pooled sensitivity and specificity by data source.**

**S4 Fig. Subgroup forest plot of pooled AUC by validation strategy.**

**S5 Fig. Subgroup forest plot of pooled AUC by data source.**

**S1 File. Extracted 2×2 contingency tables and slide-level AUC values used in all meta-analyses.**

**S2 File. Stata 17 analysis code reproducing all meta-analyses reported in this article.**

## Data availability statement

All data underlying the findings of this systematic review are provided in full within the manuscript and its Supporting information files. The Supporting information files comprise the PRISMA 2020 checklist, the review protocol registered in PROSPERO, the full search strategies, the list of all records screened with reasons for exclusion, the study characteristics table, the extracted 2×2 contingency tables and slide-level AUC values used in the analyses, the QUADAS-2 and GRADE assessments, the cohort overlap assessment, and the Stata code used to reproduce all analyses. No individual-participant data were used, and no datasets were generated that are subject to access restrictions.

